# Automated Assessment of Hypernasality in Motor Speech Disorders Using Nasal Cognate Distinctiveness and Nasal Posterior Probability

**DOI:** 10.64898/2026.09.19.26363478

**Authors:** Fenqi Wang, Rene L. Utianski, Leland R. Barnard, Joseph R. Duffy, Hugo Botha

## Abstract

**Purpose:** To evaluate whether two automated measures of nasality, Nasal Cognate Distinctiveness (NCD) and nasal posterior probability (NPP), provide clinically meaningful markers of hypernasality in speakers with motor speech disorders (MSDs) and to determine whether hypernasality-related information extends beyond stop consonants, which are typically targeted in cognate-based approaches.

**Method:** Sentence-level speech recordings were obtained from 561 patients, including 374 neurologically typical controls and 187 individuals with MSDs. NCD was computed for voiceless stops (/p/, /t/, /k/) using likelihood ratios between stops and their nasal cognates. NPP was extracted using a phonological feature-based neural network model and analyzed for stops, consonants, and vowels. Hypernasality severity was rated by speech-language pathologists. Group differences and associations with hypernasality severity were evaluated using linear mixed-effects models and Spearman correlations, and receiver operating characteristic (ROC) analyses were used to evaluate discrimination of absent versus present hypernasality. An additional 20 controls with repeat recordings were used to evaluate test-retest reliability and minimal detectable change (MDC).

**Results:** NCD generally decreased and NPP increased with increasing hypernasality severity, with phoneme-specific severity effects observed for NCD. Across stops, NCD and NPP were strongly inversely correlated and showed comparable discrimination of absent versus present hypernasality (AUCs = 0.796 and 0.730, respectively). Consonant and vowel NPP measures were both positively associated with hypernasality severity and demonstrated identical classification performance (AUC = 0.853). No significant differences in ROC performance were observed between NCD and NPP or between consonant and vowel NPP measures. Test-retest change exceeded the MDC threshold in 2 of 20 controls for NCD and 1 of 20 for NPP.

**Conclusions:** Both NCD and NPP provide objective markers of hypernasality in speakers with MSDs. The measures capture related aspects of nasalization and demonstrate comparable ability to identify clinically meaningful hypernasality. NPP captures hypernasality-related acoustic information across both consonants and vowels, with similar relationships to severity and classification performance across sound classes. Test-retest findings provide preliminary MDC benchmarks for interpreting longitudinal change.

## 1 Introduction

Hypernasality is a common speech characteristic in individuals with motor speech disorders (MSDs) and is typically associated with impaired velopharyngeal function (Duffy, 2020; Kummer, 2018). Although clinicians routinely evaluate hypernasality as part of a comprehensive motor speech assessment, objective measurement remains challenging because nasal resonance arises from complex interactions among articulatory, acoustic, and perceptual factors (Kent & Kim, 2003; Kummer, 2018). Recent advances in automatic speech processing have created opportunities to quantify hypernasality directly from speech recordings, potentially providing scalable and objective measures for clinical and research applications (Golabbakhsh et al., 2017; He et al., 2014). The present study examines two automated measures of nasalization, Nasal Cognate Distinctiveness (NCD) (Saxon et al., 2019) and nasal posterior probability (NPP) (Vásquez-Correa et al., 2019), and evaluates their relationships with clinician-rated hypernasality severity in a large cohort of speakers with MSDs.

### 1.1 Hypernasality in Motor Speech Disorders

Hypernasality is a perceptual speech characteristic resulting from excessive nasal resonance during speech production (Kummer, 2018). It occurs when velopharyngeal closure is insufficient to appropriately separate the oral and nasal cavities during speech, allowing excessive acoustic energy to be transmitted through the nasal tract (Kummer & Lee, 1996). Hypernasality can occur across dysarthria types but is most commonly associated with flaccid dysarthria, followed by spastic and hypokinetic dysarthria; it may also occur in hyperkinetic and ataxic dysarthria (Duffy, 2020). Excessive nasal resonance and excessive airflow through the nasal cavity during consonants requiring intraoral pressure can reduce speech intelligibility, alter speech naturalness, and negatively affect communicative effectiveness, making its assessment an important component of clinical motor speech evaluation (Duffy, 2020). Excessive nasal airflow during production of pressure consonants may also be perceived as audible nasal emission, which can co-occur with hypernasality but represents a perceptually distinct speech feature (Oren et al., 2020).

The acoustic manifestations of hypernasality are complex and highly variable. The percept reflects interactions among oral, nasal, and pharyngeal resonances, and its expression may differ across speakers, phonetic contexts, and underlying neurological disorders (Kummer, 2018; Stevens, 2000). Hypernasality also frequently co-occurs with other speech impairments, including articulatory imprecision, altered voice quality, and prosodic abnormalities, making it difficult to isolate acoustically (Duffy, 2020). Consequently, the relationship between perceived hypernasality and objective acoustic measurements is often nonlinear and influenced by multiple factors beyond velopharyngeal function alone (Bettens et al., 2018; Kent et al., 1989; Saxon et al., 2019).

### 1.2 Current Approaches to Hypernasality Assessment

Auditory-perceptual evaluation by speech-language pathologists remains the clinical standard for assessing hypernasality (Kummer, 2018; Kummer & Lee, 1996). Although perceptual judgments provide clinically meaningful information, they are inherently subjective and may be influenced by listener experience, rating procedures, and co-occurring speech characteristics (Brunnegård et al., 2012; Oliveira et al., 2016). Reliable perceptual assessment often requires multiple raters or extensive training, highlighting challenges related to inter-rater consistency and reproducibility (Bettens et al., 2018; Paal et al., 2005).

Several instrumental methods have been developed to supplement perceptual evaluation. Imaging techniques such as videofluoroscopy and nasoendoscopy provide direct information regarding velopharyngeal function but require specialized equipment and may be invasive or impractical for routine monitoring (Guyton et al., 2018; Stelck et al., 2011). Nasometry offers a noninvasive estimate of nasal acoustic energy and demonstrates moderate associations with perceptual judgments; however, it likewise requires dedicated instrumentation and clinical expertise for administration and interpretation (Liu et al., 2022). As a result, objective measures that can be derived directly from speech recordings remain attractive alternatives for both clinical and research applications.

Acoustic approaches to hypernasality assessment have traditionally focused on spectral characteristics associated with nasalization, including changes in formant structure, bandwidth, spectral tilt, and other resonance-related measures. Prior studies have reported associations between hypernasality and increased formant bandwidths, nasal formants and anti-formants, and changes in spectral energy distribution and cepstral characteristics (Kataoka et al., 2001; Tsai et al., 2012; Wang et al., 2026). Other investigations have shown that combinations of spectral and voice quality features can successfully distinguish hypernasal from non-hypernasal speech using statistical and machine learning approaches (Golabbakhsh et al., 2017; He et al., 2014). These findings support the feasibility of acoustic assessment while also highlighting the diversity of acoustic manifestations associated with hypernasality. Although traditional acoustic measures have demonstrated utility, the acoustic consequences of hypernasality are multifaceted and may not be adequately captured by a small set of predefined acoustic features. Advances in automatic speech processing have therefore motivated the development of more comprehensive, data-driven approaches that leverage information distributed throughout the speech signal.

### 1.3 Automated Acoustic Measures of Nasalization

Recent automated approaches to hypernasality assessment can be broadly categorized into two groups. The first uses machine learning models trained to predict hypernasality directly from acoustic features or learned speech representations. These approaches have employed cepstral features, vocal tract features, spectral representations, and deep neural network embeddings to estimate hypernasality severity or classify speakers as hypernasal or non-hypernasal (Kothadia et al., 2025; Mathad et al., 2021). While such methods often achieve strong predictive performance, the resulting measures are typically optimized for classification and may provide limited insight into the specific acoustic characteristics underlying the prediction. The second group focuses on interpretable measures that quantify acoustic correlates of nasalization directly from speech. These measures are designed to capture changes in speech production associated with increased nasal resonance and may offer greater transparency for clinical interpretation. NCD and NPP belong to this latter category (Saxon et al., 2019; Vásquez-Correa et al., 2019).

NCD is motivated by the observation that incomplete velopharyngeal closure can cause oral stops to sound more similar to nasal consonants sharing the same place of articulation (Saxon et al., 2019). For example, bilabial stops may become more similar to /m/, alveolar stops to /n/, and velar stops to /ŋ/. NCD quantifies this phenomenon using likelihood ratios derived from an acoustic model trained on healthy speech, measuring the acoustic distinctiveness between a target stop and its nasal cognate (Saxon et al., 2019). Lower NCD values indicate reduced oral–nasal distinctiveness and therefore greater evidence of nasalization. This approach is grounded in the phonetic observation that nasalization can reduce acoustic contrasts between oral consonants and their nasal counterparts by introducing nasal resonance and anti-resonances into the speech signal (Stevens, 2000).

A complementary strategy is provided by phonological feature-based neural network models such as Phonet (Vásquez-Correa et al., 2019). Rather than evaluating contrasts between specific phonemes, Phonet estimates posterior probabilities for phonological feature classes directly from continuous speech. In the present study, NPP represents the likelihood that a speech segment exhibits acoustic characteristics associated with the nasal class. Because NPP is computed at the frame level and is not restricted to specific stop–nasal contrasts, it may provide a broader characterization of nasal resonance that can be evaluated across multiple phonetic contexts, including both consonants and vowels (Cernak et al., 2017; Vásquez-Correa et al., 2019).

Despite the promise of these approaches, several questions remain unresolved. First, relatively little work has directly compared stop-specific and phonological feature-based measures of hypernasality within the same clinical population. Second, because NCD is restricted to stop consonants, it remains unclear whether automated measures of nasalization can be extended to broader classes of speech sounds. Hypernasality is fundamentally a resonance phenomenon that affects speech globally, suggesting that acoustic markers may be detectable beyond the stop consonants traditionally examined in the literature (Kummer, 2018; Stevens, 2000). Finally, because hypernasality is often considered perceptually most salient in vowels, vowels may be particularly sensitive to resonance-related changes (Pereira & Sell, 2024). However, relatively little is known about whether consonants and vowels provide comparable information about hypernasality severity. Addressing these questions may help clarify how hypernasality manifests across different speech sounds and inform the development of more flexible approaches to automated assessment.

### 1.4 The Current Study

The present study evaluated two automated measures of hypernasality in a large cohort of speakers with MSDs and neurologically typical controls. Specifically, we examined NCD, a stop-specific measure of nasalization, and NPP, a phonological feature-based measure of nasal resonance.

Three aims were addressed. First, we examined whether NCD and NPP differentiated speakers with MSDs from matched controls. Second, we evaluated the relationship between NCD and NPP across voiceless stops (/p/, /t/, and /k/) to determine whether the two measures captured similar aspects of nasalization. Third, we examined whether hypernasality-related information extends beyond stop consonants by comparing NPP measures derived from consonants and vowels. This analysis was intended to determine whether broader classes of speech sounds capture similar relationships with hypernasality severity and whether consonants and vowels differ in their utility as markers of hypernasality.

We hypothesized that the presence of hypernasality would be associated with lower NCD values and higher NPP values. We further expected both measures to correlate with perceptual ratings of hypernasality and to demonstrate good discrimination between speakers with and without clinically perceived hypernasality. Finally, because hypernasality may be particularly salient perceptually in vowels, differences between consonant and vowel NPP were of clinical interest; however, the comparison was treated as exploratory and no directional a priori hypothesis regarding relative sensitivity was specified.

## 2 Methods

### 2.1 Dataset

The dataset included sentence-level speech recordings from 561 patients seen at Mayo Clinic, comprising 374 neurologically typical controls and 187 individuals diagnosed with MSDs. Speech samples were collected through a web-based, self-administered speech protocol, which includes a sentence repetition task. Patients were instructed to read aloud the sentence “*My physician wrote out a prescription*” using their natural, everyday speaking voice. The sentence contains two /p/ tokens in *prescription* (one syllable-initial and one syllable-final before /ʃ/; only syllable-initial /p/ used for comparing NCD with NPP), two /t/ tokens in *wrote* and *out* (both syllable-final), and one /k/ token within the consonant cluster in *prescription*. The sentence was selected because it contains the voiceless stops required for NCD estimation and includes both consonantal and vocalic contexts for NPP analysis. Although a single sentence limits phonetic sampling, it provides a standardized speech context across speakers and supports direct comparison of automated measures under uniform recording conditions.

Patients were not excluded on the basis of co-occurring MSD subtypes; MSD diagnostic subtype distributions were as follows: phonetic predominant apraxia of speech (*n* = 6), prosodic predominant apraxia of speech (*n* = 13), ataxic dysarthria (*n* = 45), flaccid dysarthria (*n* = 39), hyperkinetic dysarthria (*n* = 49), hypokinetic dysarthria (*n* = 29), and spastic dysarthria (*n* = 40), with 32 patients receiving two diagnoses and 1 patient receiving three diagnoses. An additional 20 neurologically typical controls completed the same speech protocol at two time points (> 90 days apart) to assess test–retest reliability and minimal detectable change (MDC) for NCD and NPP for the nasal cognate stops (/p/, /t/, and /k/). These latter participants were included only in the reliability and MDC analyses. All patients provided written informed consent, and the study was approved by the Mayo Clinic Institutional Review Board.

### 2.2 Automated Nasality Measures

#### 2.2.1 Nasal Cognate Distinctiveness

Nasal Cognate Distinctiveness (NCD) was used to derive phoneme-level measures of nasalization at voiceless stops. NCD quantifies the degree to which an observed acoustic segment sounds more like a plosive or its corresponding nasal cognate, computed as a log likelihood ratio between the two (Saxon et al., 2019)^1^:

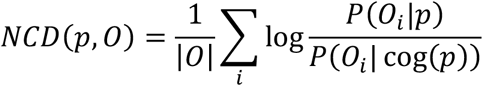

where O is the acoustic observation with frame count |O|, p is the transcript-determined phoneme, and cog(p) is a cognate mapping function that maps voiceless stops to their corresponding nasal cognates at the same place of articulation: /p/ → /m/ (bilabial), /t/ → /n/ (alveolar), and /k/ → /ŋ/ (velar). Probabilities were evaluated using Viterbi alignments from a triphone Gaussian Mixture Model–Hidden Markov Model (GMM-HMM) acoustic model trained on 960 hours of healthy native English speech from the LibriSpeech corpus (Panayotov et al., 2015). Higher NCD values reflect greater distinctiveness between the intended stop and its nasal cognate, as expected in healthy speech, whereas lower or negative values indicate that the stop sounds more like its nasal cognate, consistent with hypernasality. NCD scores were computed for each phoneme token and z-normalized using the matched non-MSD control group:

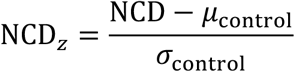

where *μ_control_* and *σ_control_* are the mean and standard deviation of NCD scores in the matched non-MSD control group

#### 2.2.2 Nasal Posterior Probability

To complement NCD, nasal posterior probabilities (NPP) were extracted using Phonet (Vásquez-Correa et al., 2019)^2^, a neural network framework that estimates the posterior probability of phonological feature classes directly from continuous speech. Phonet can generate posterior probabilities for a user-specified set of phonological classes; in the present study, nasal posterior probabilities were derived by evaluating the likelihood that each speech frame belongs to the nasal phonological class, defined by the nasal set /m, n, ŋ/. Unlike NCD, which evaluates stop-specific nasalization via likelihood ratios between a target plosive and its nasal cognate, NPP provides a more general characterization of nasal resonance at the frame level without requiring phoneme-specific cognate comparisons or transcript alignment.

Audio recordings were processed using the English-adapted Phonet framework. The model generated frame-level posterior probabilities of nasal manner of articulation, which were aggregated within phoneme boundaries obtained from forced alignment. To minimize the influence of forced alignment boundary artifacts, posterior probability averaging was restricted to the central frames of each phoneme segment rather than the full segment duration (Tang et al., 2023). For a segment containing *N* frames, posterior probabilities were averaged over the retained frames:

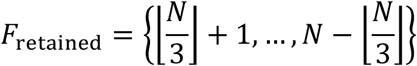

where *N* denotes the total number of frames in the segment. Thus, approximately one-third of the frames were excluded from both the onset and offset, retaining only the temporally stable central portion. For example, a 9-frame segment retains frames 4–6, whereas a 12-frame segment retains frames 5–8.

NPP values were computed separately for three phonological contexts: (1) voiceless stops (/p/, /t/, /k/), paralleling the NCD analysis; (2) all consonants; and (3) vowels. Prior to analysis, raw NPP values were transformed to logit units and z-normalized using the same matched non-MSD control group used for NCD normalization:

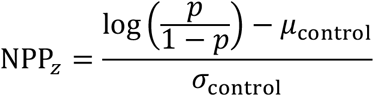

where *p* is the raw nasal posterior probability and *μ_control_* and *σ_control_* are the mean and standard deviation of logit-transformed NPP values in the matched non-MSD control group. Higher NPP values reflect greater nasal resonance and are expected to increase with hypernasality severity.

### 2.3 Diagnosis and Hypernasality Measure

MSD diagnoses and ratings of hypernasality were established by board-certified speech-language pathologists with expertise in neurogenic communication disorders at Mayo Clinic. Diagnostic classification was based on auditory-perceptual evaluation of a standardized speech-language assessment battery, including repetition, reading, and spontaneous speech tasks. Patients with MSD were classified into diagnostic groups based on their predominant subtype presentation.

Hypernasality was rated at the sentence level using a six-level ordinal scale: normal (1), equivocal (1.5), mild (2), moderate (3), marked (4), and severe (5), reflecting the overall degree of perceived nasal resonance across the entire sentence recording. Transcripts for each recording were generated using Whisper (large), an automatic speech recognition model (Radford et al., 2023), by comparing the Whisper-generated transcripts against the reference sentence. Word error rate (WER) was computed as a measure of speech intelligibility, with higher values reflecting greater deviation from the intended utterance. Recordings with WER ≥ 1.0 were excluded to ensure adequate speech quality for acoustic analysis (with the following hypernasality ratings: normal (*n* = 20), equivocal (*n* = 2), mild (*n* = 4), and moderate (*n* = 5)). Following WER filtering, no speakers retained in the final sample were rated as Marked or Severe on the hypernasality scale, and hypernasality severity was distributed as follows among MSD patients: normal (*n* = 155), equivocal (*n* = 7), mild (*n* = 12), and moderate (*n* = 12), and marked (*n* = 1).

### 2.4 Statistical Analysis

All statistical analyses were conducted in R (R Core Team, 2026). Propensity score matching was performed at the subject level using nearest-neighbor matching with a 2:1 control-to-MSD ratio, with age and gender as matching covariates, using the MatchIt package (Ho et al., 2011).

Differences between controls and speakers with MSD across hypernasality severity levels were evaluated using linear mixed-effects models (LMEMs), implemented using the lmerTest package (Kuznetsova et al., 2017). For voiceless stops, separate models were fitted for NCD and NPP with severity group, phoneme, and their interactions as fixed effects and subject as a random intercept. Severity group comprised controls and MSD speakers categorized by clinician-rated hypernasality severity. For the consonant–vowel analyses, analogous models included severity group, phone class, and their interaction as fixed effects.

Control-referenced post hoc comparisons were performed using estimated marginal means with the emmeans package (Lenth & Piaskowski, 2025). Treatment-versus-control contrasts were evaluated separately by phoneme or phone class, with multivariate-*t* adjustment for multiple comparisons.

To evaluate changes associated with increasing hypernasality severity within speakers with MSD, additional LMEMs were fitted after excluding controls. For the voiceless-stop analyses, hypernasality severity, phoneme, and their interaction were included as fixed effects, with subject as a random intercept (see formula 1). Linear severity trends were evaluated using contrast weights reflecting the spacing of the hypernasality scale, with Holm adjustment across phoneme-specific trend tests. Pairwise comparisons among phoneme-specific linear trends were adjusted using Tukey’s method, and consecutive severity-level contrasts were evaluated with Holm adjustment for multiple comparisons. For the consonant and vowel analysis, an analogous MSD-only model was fitted with hypernasality severity, phone class, and their interaction (see formula 2); an overall linear severity trend averaged across phone classes was evaluated when the interaction was not significant.

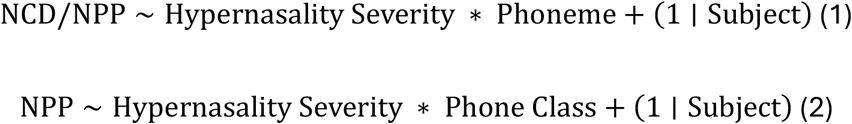

Spearman rank correlations were computed to examine associations between NCD, NPP, and hypernasality severity ratings, both across all MSD speakers and after restricting analyses to speakers with abnormal hypernasality (rating ≥ 2). Stop-level correlations were examined across all stops and separately by phoneme, and associations between NPP and hypernasality severity were additionally examined separately for consonants and vowels, with the asymptotic approximation used to accommodate tied values.

MDC was evaluated for NCD and NPP at two time points for the nasal cognate stops from the 20 newly recruited control participants. Prior to the MDC analysis, z-scores were applied back to the token-level NCD and NPP data using the corresponding subject-level mean and standard deviation. For each measure, the standard error of measurement (SEM) was estimated from the standard deviation of the *z*-normalized measure at the first time point:

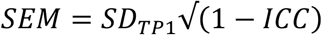

The MDC at the 95% confidence level was then calculated as:

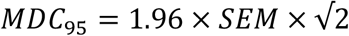

where 1.96 corresponds to the 95% confidence level and √2 accounts for measurement error associated with both time points. Test–retest reliability was assessed using intraclass correlation coefficients (ICCs) based on the two measurements obtained from the 20 control participants. Single-measure, absolute-agreement ICCs were used for the MDC calculations and were 0.14 for NCD and 0.29 for NPP (Koo & Li, 2016). For each participant, longitudinal change was calculated as:

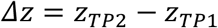

where Δz represents the change in the z-normalized score from TP1 to TP2, with positive values indicating an increase and negative values indicating a decrease over time. The absolute magnitude of this change was compared with the corresponding MDC_95_; values exceeding the MDC threshold were considered to represent change beyond that expected from measurement error.

Receiver operating characteristic (ROC) analyses were conducted to evaluate the ability of NCD and NPP to discriminate between normal (i.e., normal and equivocal) and abnormal (i.e., mild and moderate) hypernasality at the subject level. Area under the ROC curve (AUC) values were computed for each measure, and pairwise comparisons between ROC curves were conducted using DeLong’s test, implemented using the pROC package (Robin et al., 2011).

## 3 Results

### 3.1 Nasal Cognate Stops

Linear mixed-effects modeling including controls revealed significant main effects of hypernasality severity group and phoneme, as well as significant hypernasality × phoneme interactions, for both NCD and NPP. Post hoc comparisons showed phoneme-specific effects for both measures. For /k/, NCD was significantly lower than controls across all hypernasality levels (see Figure 1), whereas NPP was significantly higher than controls at all hypernasality levels except the mild level (see Figure 2). No significant differences from controls were observed for /p/ or /t/ for either measure.

**Figure 1.**
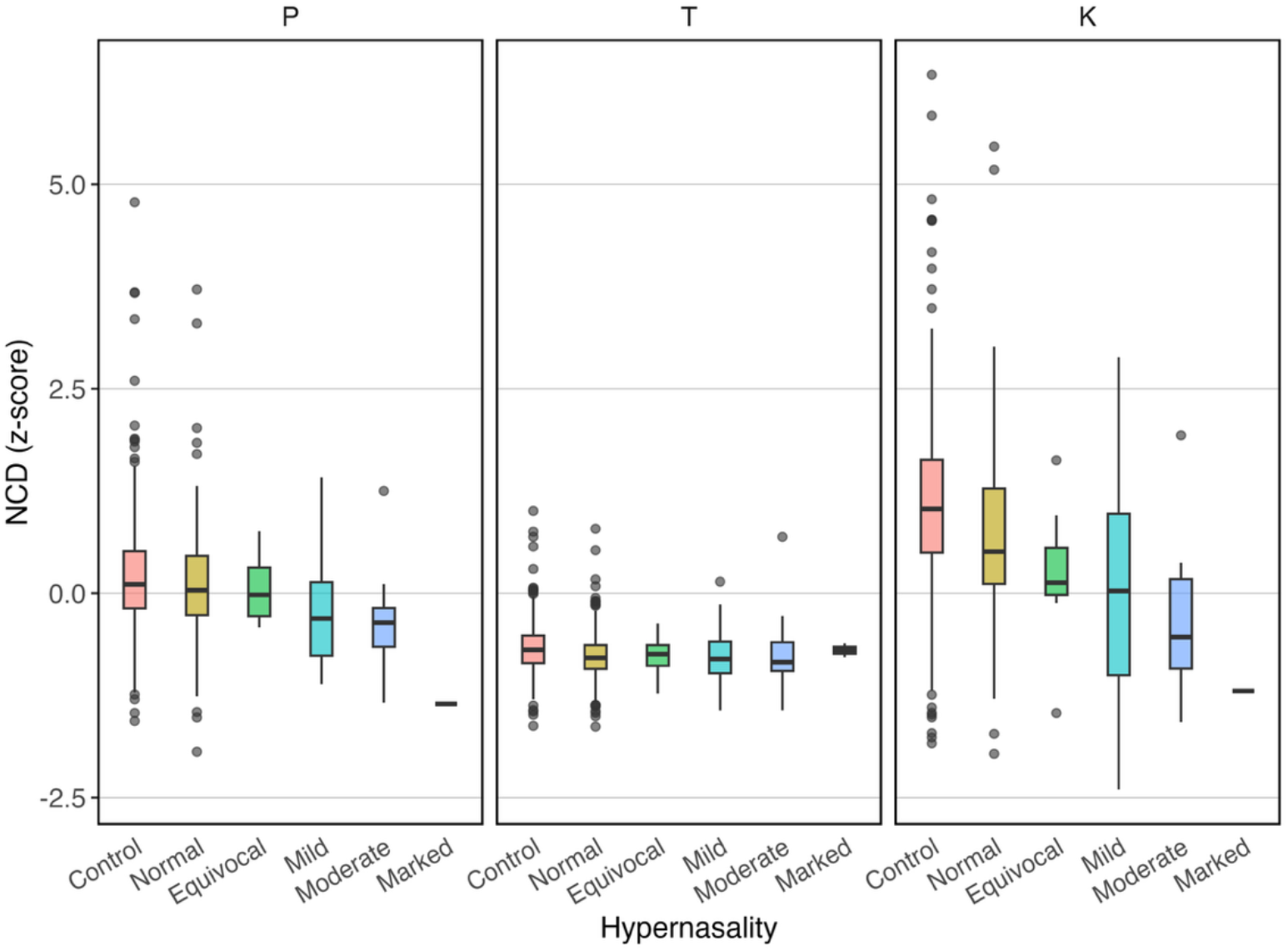
NCD (z-scores) across hypernasality severity levels by phoneme (/p/, /t/, /k/).

**Figure 2.**
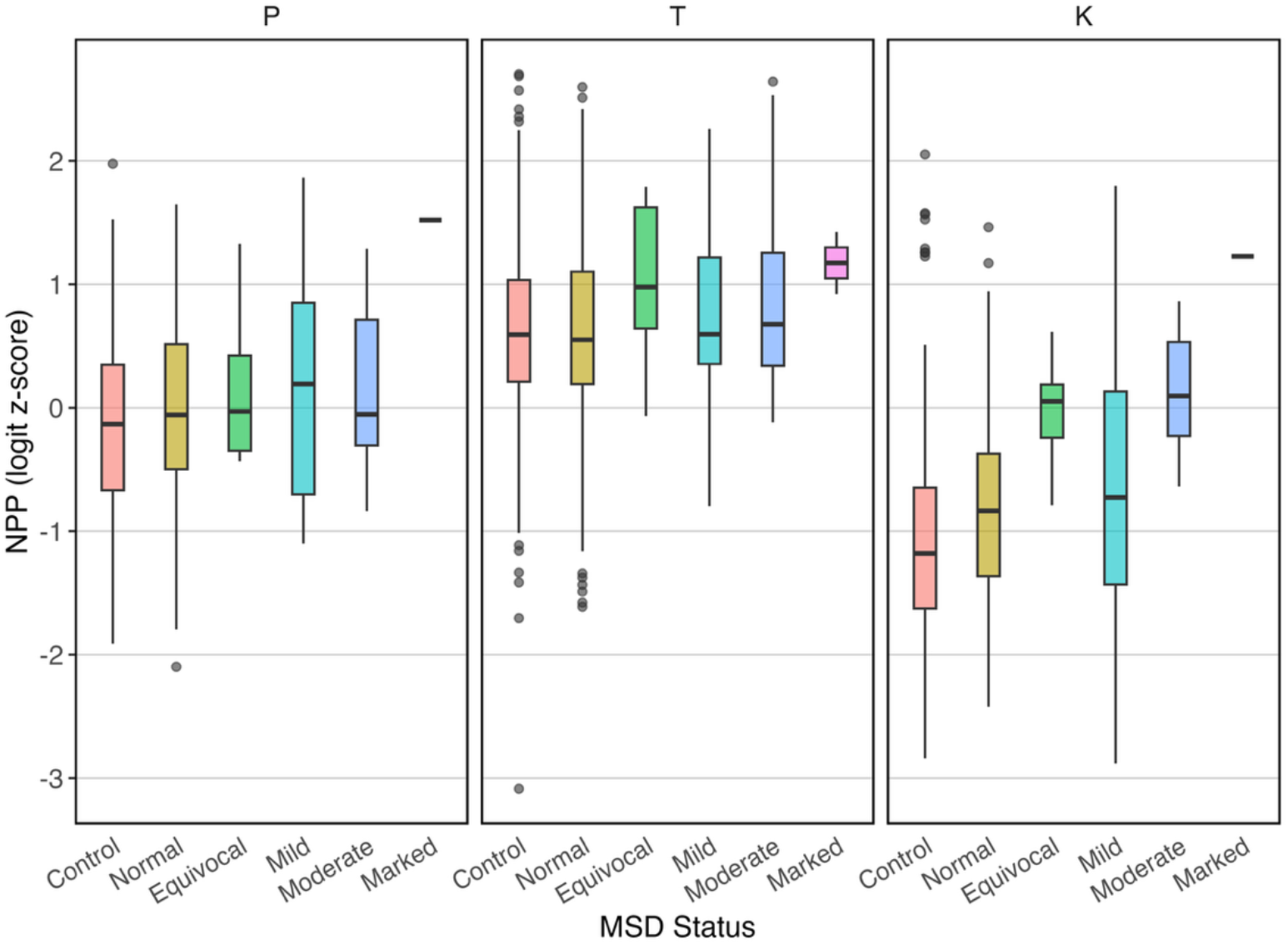
NPP (logit z-scores) across hypernasality severity levels by phoneme (/p/, /t/, /k/).

Within MSD patients, linear mixed-effects modeling revealed significant main effects of hypernasality (*F*(4, 214.75) = 8.03, *p* ff .001) and phoneme (*F*(2, 551.00) = 6.54, *p* = .002), as well as a significant hypernasality × phoneme interaction (*F*(8, 551.00) = 4.19, *p* ff .001). Holm-adjusted linear trend analyses showed a significant decrease in NCD with increasing hypernasality for /k/ (*t*(724) = −2.83, *p* = .015), but not for /p/ or /t/ (see Figure 3). Tukey-adjusted comparisons of the linear trends showed that the severity trend for /k/ differed significantly from that for /t/ (*t*(551) = 2.48, *p* = .036), whereas the /p/ trend did not differ significantly from either /t/ or /k/. No adjacent hypernasality severity levels differed significantly within any phoneme after correction for multiple comparisons. For NPP, significant main effects of hypernasality (*F*(4, 212.25) = 7.40, *p* ff .001) and phoneme (*F*(2, 551.00) = 10.45, *p* ff .001) were observed, whereas the hypernasality × phoneme interaction was not significant. Holm-adjusted linear trend analyses showed significant increases in NPP with increasing hypernasality for /k/ (*t*(717) = 2.69, *p* = .007), but not for /p/ and /t/ (see Figure 4).

**Figure 3.**
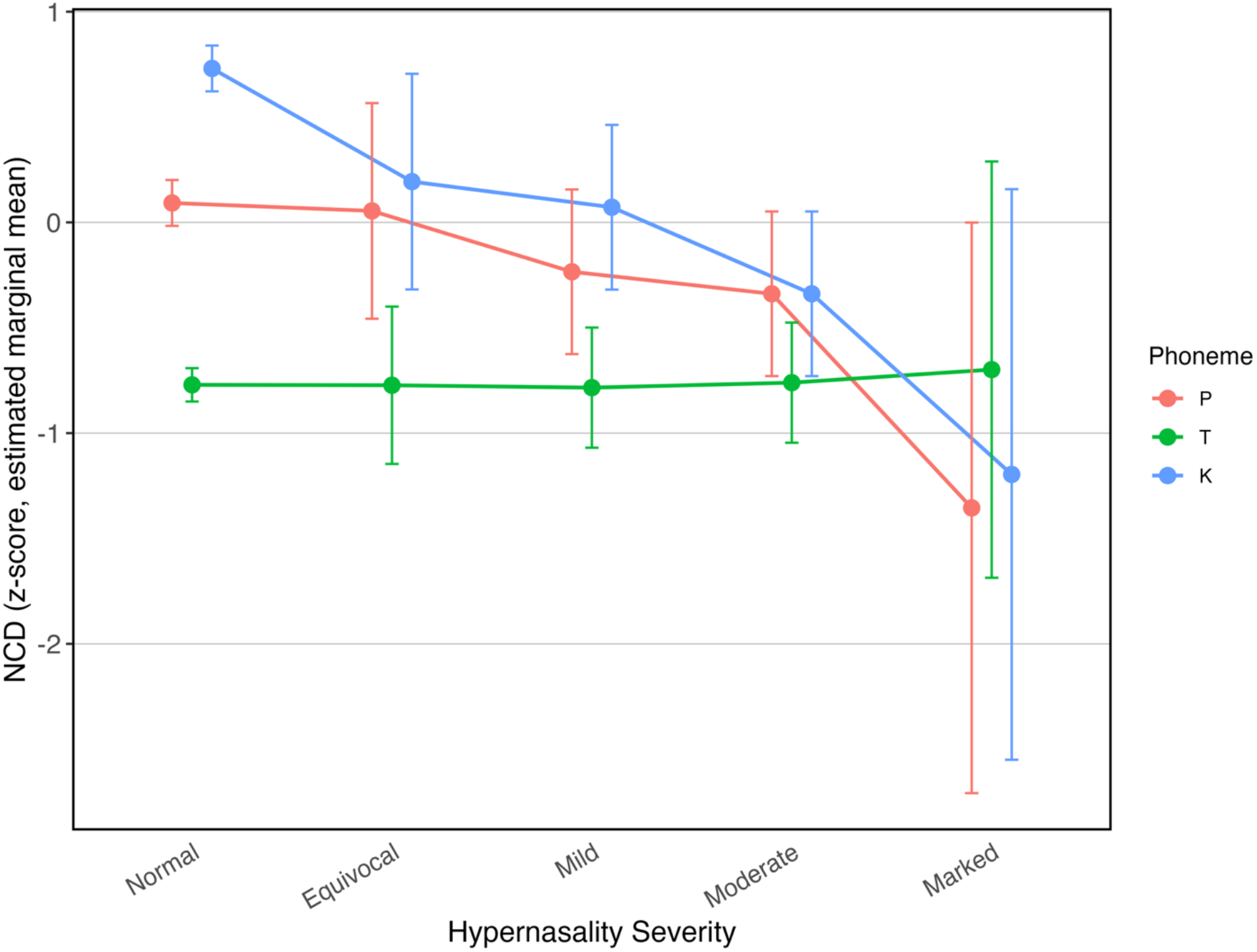
Estimated marginal means of NCD (z-scores) across hypernasality severity levels by phoneme (/p/, /t/, /k/) for MSD patients, derived from the linear mixed-effects model. Error bars represent confidence intervals.

**Figure 4.**
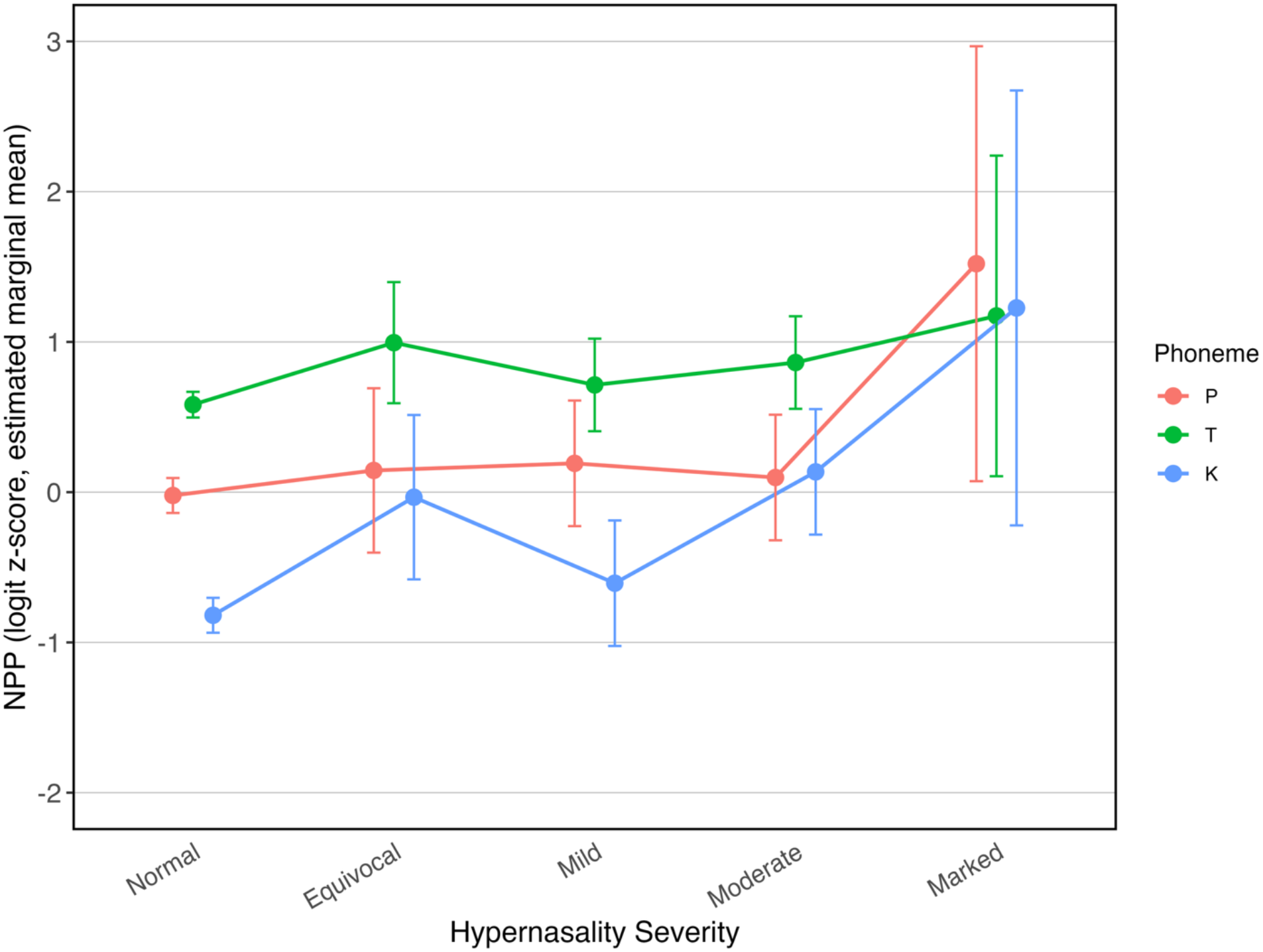
Estimated marginal means of NPP (logit z-scores) across hypernasality severity levels by phoneme (/p/, /t/, /k/) for MSD patients, derived from the linear mixed-effects model. Error bars represent confidence intervals.

Across all stops, NCD was negatively correlated with hypernasality severity (*r* = −0.179, *p* ff .001), indicating lower NCD values with increasing severity. At the phoneme level, significant negative correlations were observed for /p/ (*r* = −0.216, *p* = .003) and /k/ (*r* = −0.294, *p* ff .001), but not for /t/. In contrast, NPP was positively correlated with hypernasality severity (*r* = 0.161, *p* ff .001), with significant positive correlations observed for /t/ (*r* = 0.153, p = .036) and /k/ (*r* = 0.318, *p* ff .001). NCD and NPP were strongly negatively correlated (*r* = −0.647, *p* ff .001), with significant inverse correlations observed for each phoneme. When analyses were restricted to speakers with abnormal hypernasality (rating ≥ 2), NCD and NPP remained significantly correlated across stops (*r* = −0.423, *p* ff .001); however, the relationship between NCD and NPP was phoneme-specific. A strong negative correlation was observed for /k/ (*r* = −0.674, *p* ff .001), whereas correlations were not significant for /p/ or /t/. Similarly, hypernasality severity was significantly and positively correlated with NPP for /k/ (*r* = 0.439, *p* = .028), but not for /p/ or /t/. No significant correlations were found between hypernasality severity and NCD.

As shown in Figure 5, ROC analyses demonstrated good discrimination between normal and abnormal hypernasality for both NCD (AUC = 0.796) and NPP (AUC = 0.730). However, DeLong’s test showed no significant difference in classification performance between the two measures.

**Figure 5.**
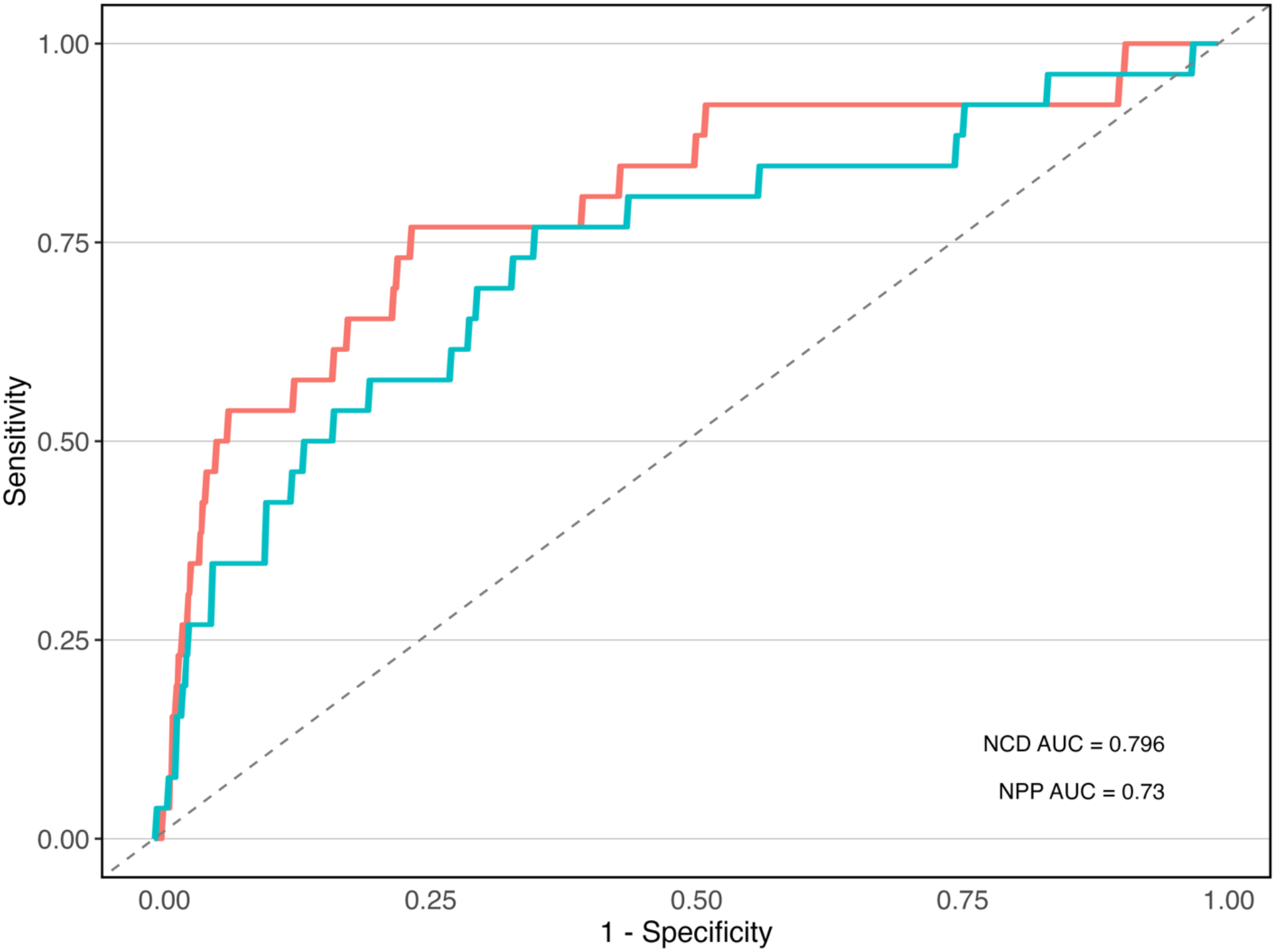
ROC curves for NCD and NPP discriminating normal versus abnormal hypernasality. AUC values are annotated in the lower right corner.

The MDC_95_ was 2.57 z-score units for NCD and 2.34 z-score units for NPP. Test–retest change exceeded these thresholds in 2 of 20 controls (10%) for NCD and 1 of 20 controls (5%) for NPP (see Figure 6).

**Figure 6.**
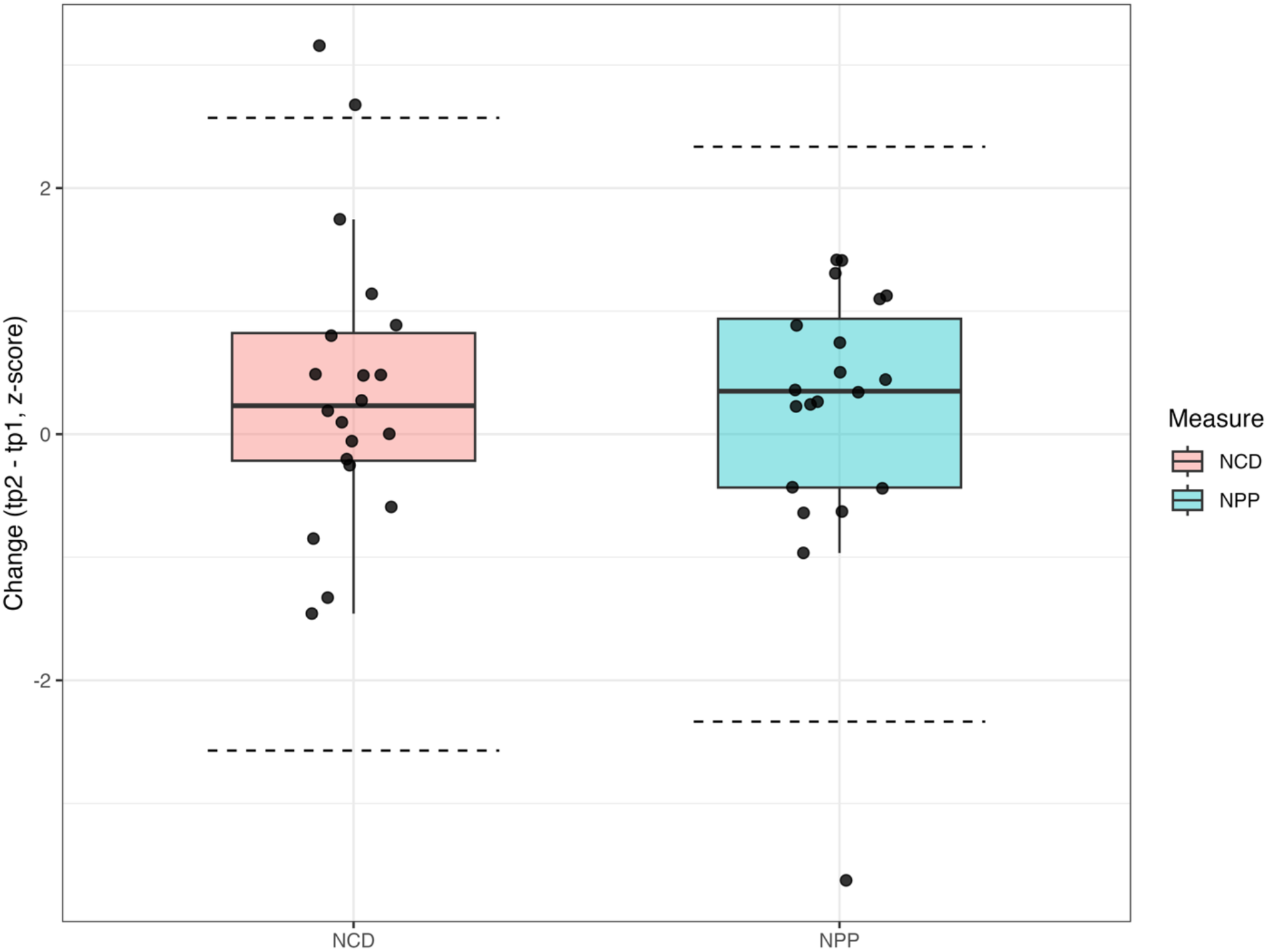
Test–retest changes in NCD and NPP relative to minimal detectable change (MDC_95_) thresholds. Dashed lines indicate ±MDC_95_ thresholds (NCD: ±2.57 z-score units; NPP: ±2.34 z-score units).

### 3.2 Consonants vs. Vowels

Linear mixed-effects modeling including controls revealed a significant main effect of hypernasality severity group (*F*(5, 606.30) = 30.87, *p* < .001), whereas neither the main effect of phone class (*F*(1, 14019.00) = 0.96, *p* = .327) nor the hypernasality severity group × phone class interaction (*F*(5, 14019.00) = 0.95, *p* = .446) was significant. Post hoc comparisons showed that consonant NPP was significantly higher than controls across all hypernasality levels. Vowel NPP was significantly higher than controls at the normal, mild, moderate, and marked levels, but not at the equivocal level (see Figure 7).

**Figure 7.**
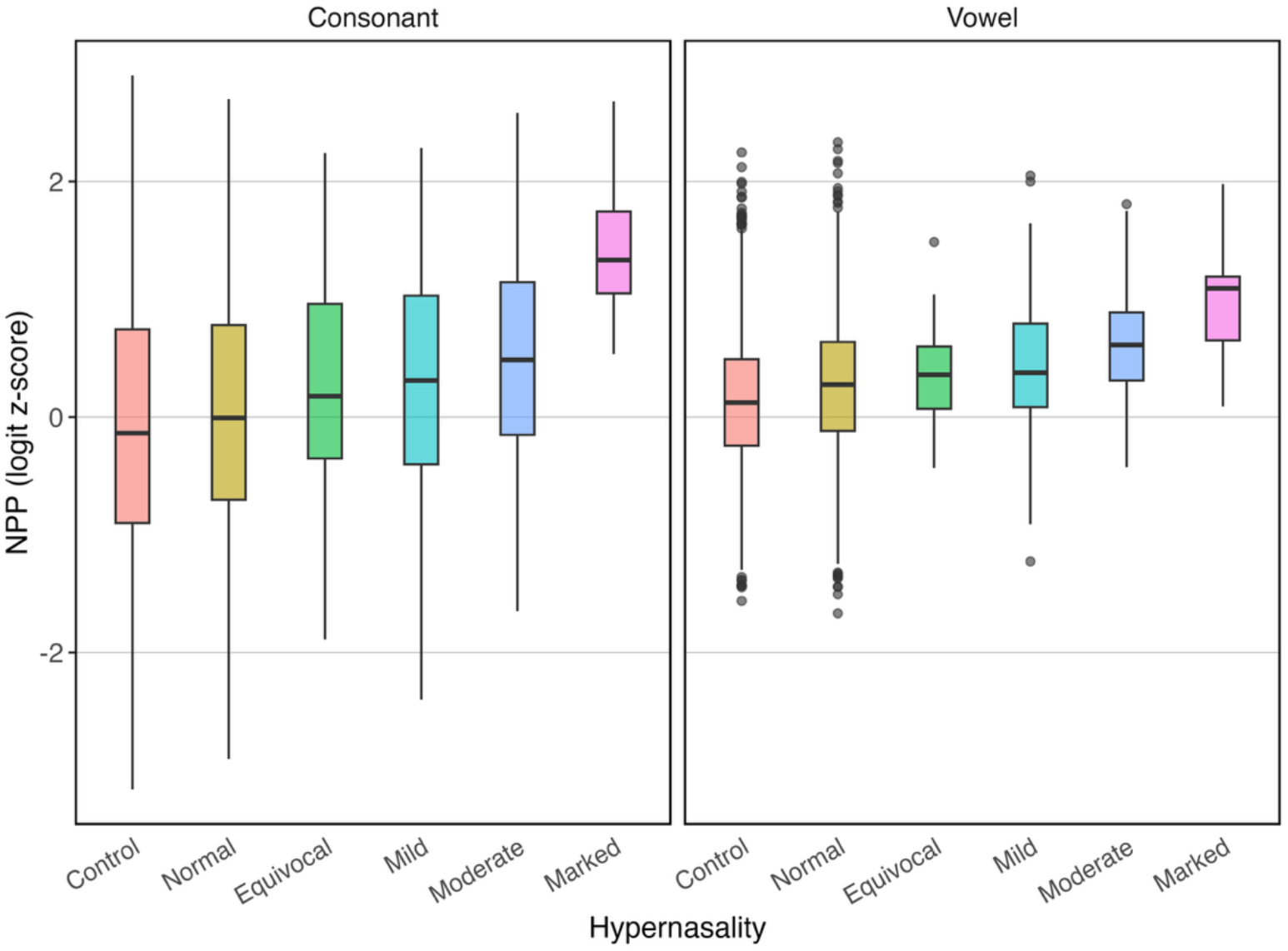
NPP (logit z-scores) across hypernasality severity levels for consonants and vowels.

Within MSD patients, a linear mixed-effects model revealed significant main effects of hypernasality (*F*(4, 193.20) = 14.49, *p* ff .001), whereas neither the main effect of phone class nor the hypernasality × phone class interaction was significant. A linear trend analysis averaged across phone classes showed that NPP increased significantly with increasing hypernasality severity (*β* = 0.333, *SE* = 0.073, *t*(193) = 4.57, *p* ff .001) (see Figure 8).

**Figure 8.**
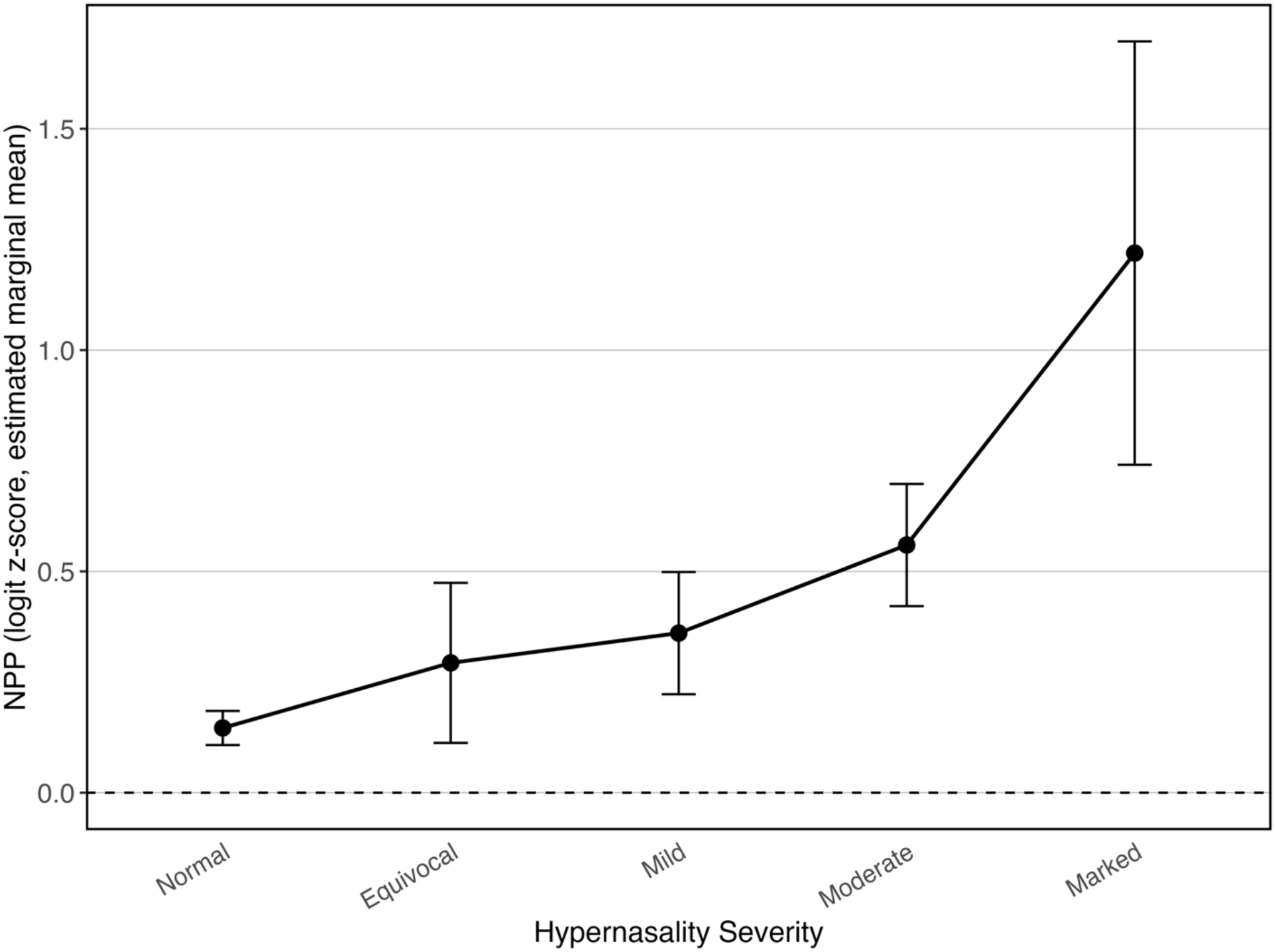
Estimated marginal means of NPP (logit z-scores) across hypernasality severity levels for MSD patients, derived from the linear mixed-effects model. Error bars represent confidence intervals.

Both consonant NPP (*r* = 0.416, *p* ff .001) and vowel NPP (*r* = 0.332, *p* ff .001) were positively associated with hypernasality severity across all MSD patients. When analyses were restricted to speakers with abnormal hypernasality (*n* = 25), the association remained significant for consonant NPP (*r* = 0.423, *p* = .035), while vowel NPP showed a similar positive association at the conventional significance threshold (*r* = 0.397, *p* = .050).

ROC analyses indicated high and identical classification accuracy for both consonant and vowel NPP (AUCs = 0.853), as presented in Figure 9. DeLong’s test revealed no difference in discriminative performance between phone classes, suggesting that consonant and vowel NPP are equally effective markers of hypernasality.

**Figure 9.**
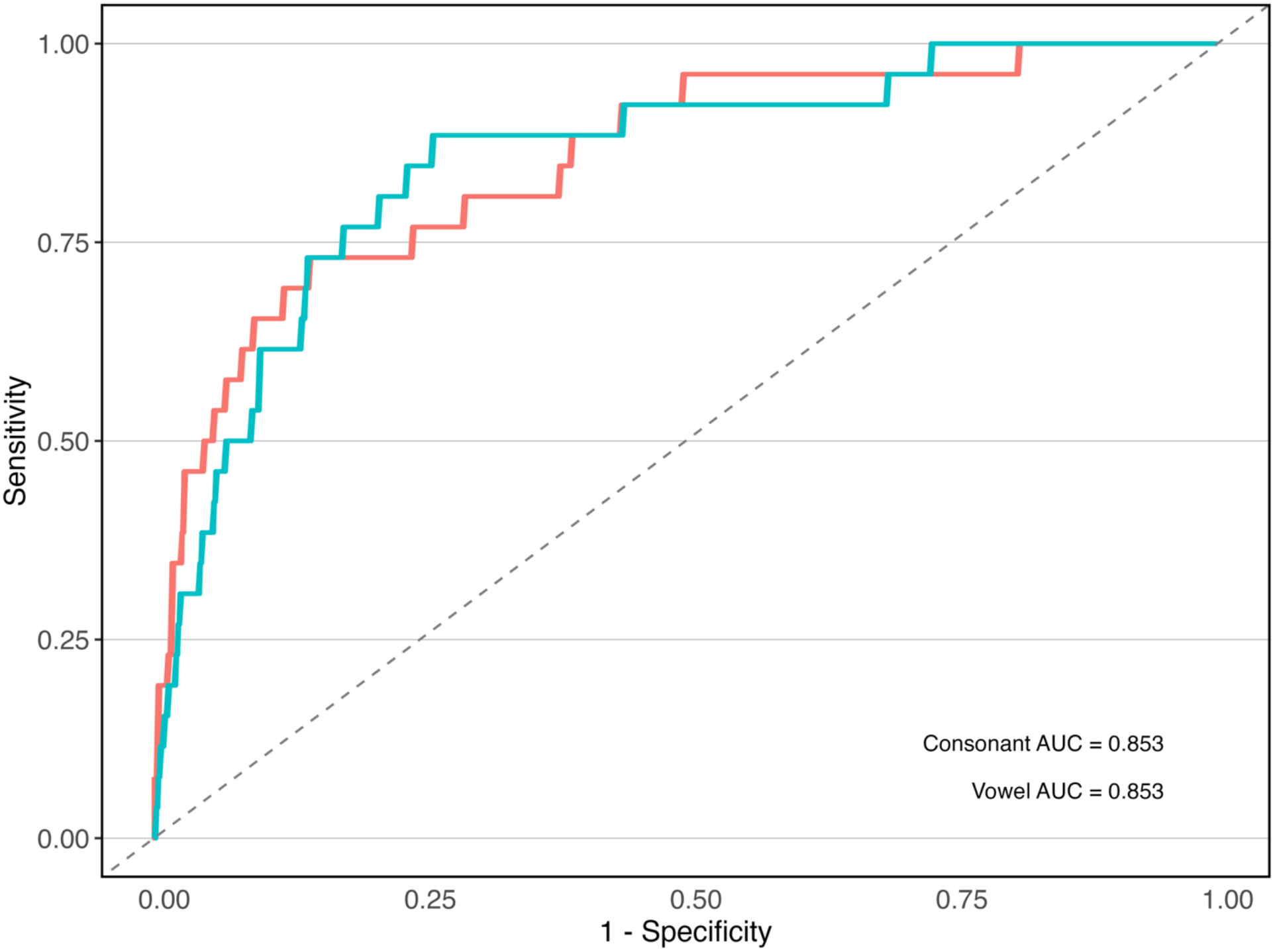
ROC curves for consonant and vowel NPP discriminating normal versus abnormal hypernasality. AUC values are annotated in the lower right corner.

## 4 Discussion

This study evaluated two automated measures of hypernasality, Nasal Cognate Distinctiveness (NCD) and nasal posterior probability (NPP), in a large cohort of speakers with MSDs. Three aims were addressed: determine whether NCD and NPP differentiates MSD speakers from controls, examine the relationship between NCD and NPP within voiceless stops, and evaluate whether hypernasality-related information extends beyond stop consonants. Overall, the findings support the utility of both measures as objective markers of hypernasality and largely support the study hypotheses.

### 4.1 Automated Measures Track Hypernasality Severity

The first aim was to determine whether NCD and NPP were associated with clinician-rated hypernasality and distinguished MSD speakers from controls. Overall, these measures show promise for identifying the presence of hypernasality and provide preliminary evidence of sensitivity to severity. Pairwise comparisons demonstrated significant differences between controls and multiple hypernasality severity levels for both measures. Overall, NCD decreased and NPP increased with increasing hypernasality severity, although these relationships varied across stop consonants. For NCD, the severity effect was phoneme-specific, with the clearest decrease observed for /k/. For NPP, an overall severity effect was observed, with /k/ showing the clearest severity-related increase. Correlation analyses similarly showed modest overall associations between both measures and hypernasality severity.

These findings support the primary hypothesis that the presence of perceived hypernasality and its severity is associated with lower NCD values and higher NPP values. The directionality of these effects is consistent with the theoretical foundations of both measures (Kummer, 2018; Stevens, 2000). As hypernasality increases, oral stops become acoustically less distinct from their nasal cognates, resulting in lower NCD values. At the same time, speech segments become increasingly likely to exhibit acoustic characteristics associated with nasal consonants, leading to higher NPP values.

Although the observed correlations with severity were statistically significant, their magnitudes were modest. The modest correlations may also reflect the restricted range of perceptual ratings: the analyzed sample was dominated by normal through moderate ratings, with only one speaker rated as marked and none rated as severe. Hypernasality is influenced by multiple acoustic and physiological factors, and perceptual judgments may also be affected by co-occurring speech impairments (Bettens et al., 2018; Duffy, 2020; Liu et al., 2022). Consequently, no single acoustic measure is likely to account for a large proportion of variance in perceptual severity ratings. Nevertheless, the consistent relationships observed across multiple analyses indicate that both NCD and NPP capture clinically relevant aspects of nasal resonance (Golabbakhsh et al., 2017; Mathad et al., 2021).

### 4.2 NCD and NPP Capture Similar Aspects of Nasalization

The second aim was to evaluate the relationship between NCD and NPP across voiceless stops. The results strongly suggest that the two measures capture related aspects of stop nasalization. Across all stop consonants, NCD and NPP exhibited a substantial negative correlation, indicating that stops judged by the acoustic model to be less distinct from their nasal cognates also received higher nasal posterior probabilities. Importantly, both measures were associated with hypernasality severity overall and demonstrated comparable classification performance in ROC analyses, although their phoneme-specific relationships with hypernasality severity differed slightly.

Despite being derived from different computational frameworks, both measures appear sensitive to the same underlying phenomenon: acoustic consequence of abnormal oral-nasal coupling, whereby incomplete velopharyngeal closure increases acoustic interaction between the oral and nasal cavities during speech (Oren et al., 2020; Watterson, 2020).

NCD approaches this problem through phoneme-specific comparisons between oral stops and their nasal cognates, whereas NPP estimates the probability that acoustic frames exhibit nasal characteristics (Cernak et al., 2017; Stevens, 2000). The strong inverse relationship between the two measures suggests that they provide converging evidence of nasalization-related acoustic change rather than capturing entirely distinct aspects of speech production (Kent, 1996; Stevens, 2000).

Notably, neither measure demonstrated superior classification performance. Although NCD produced a numerically higher AUC than NPP, the difference was not statistically significant. This result suggests that a more targeted stop-specific measure does not necessarily provide greater sensitivity to hypernasality than a broader phonological feature-based approach. From a practical perspective, this finding is encouraging because NPP can be computed across a wider range of speech sounds and does not require explicit stop–nasal cognate comparisons.

### 4.3 Hypernasality Extends Beyond Stop Consonants

The third aim was to determine whether hypernasality-related information extends beyond stop consonants by examining NPP measures derived from consonants and vowels. The results indicate that both consonant and vowel NPP values were positively associated with hypernasality severity and demonstrated equivalent classification performance. NPP increased significantly with increasing severity across phone classes, with no evidence that this relationship differed between consonants and vowels, and both measures achieved identical ROC performance when distinguishing speakers with and without abnormal hypernasality.

These findings demonstrate that NPP captures hypernasality-related acoustic information across broader classes of speech sounds, consistent with the established physiological basis of hypernasality as a resonance disorder that affects speech beyond stop consonants (Kuehn & Moller, 2000; Kummer, 2018). Unlike articulatory distortions that may be confined to specific phonemes, altered velopharyngeal function affects the acoustic properties of speech more globally (Duffy, 2020). Consequently, markers of nasal resonance can be observed in both consonants and vowels (Chen, 1997; Pereira & Sell, 2024).

Notably, overall NPP did not differ significantly between consonants and vowels, and there was no evidence that the relationship between NPP and hypernasality severity differed between phone classes. At the same time, the absence of meaningful differences between the two classes suggests that automated assessment of hypernasality may not need to focus exclusively on vowels or on consonants traditionally associated with nasalization.

### 4.4 Clinical and Methodological Implications

Several implications emerge from these findings. First, the results provide further evidence that automated acoustic measures can capture clinically meaningful information about hypernasality directly from speech recordings (Golabbakhsh et al., 2017; Kothadia et al., 2025; Mathad et al., 2021). Such measures may complement perceptual assessment by providing objective and reproducible indices of nasal resonance abnormalities that are not dependent on listener judgment. The MDC analysis further showed that test–retest changes remained within the MDC thresholds for most controls, with only 10% for NCD and 5% for NPP exceeding the respective thresholds. These findings provide preliminary thresholds for distinguishing longitudinal change from measurement error, although further validation in larger samples is warranted.

Second, the comparable performance of NCD and NPP suggests that hypernasality can be quantified using either targeted phoneme-specific analyses or broader phonological feature-based approaches, although the observed phoneme-specific differences indicate that targeted measures may be sensitive to phonetic context. While NCD offers a theoretically interpretable measure grounded in oral–nasal contrasts, NPP may be more flexible because it can be computed across a wider range of speech sounds and does not depend on specific phonetic environments.

Finally, the consonant–vowel analyses show that NPP captures hypernasality-related information across multiple sound classes. This finding supports the use of more comprehensive speech sampling strategies when developing automated hypernasality measures and suggests that future systems may benefit from integrating information across diverse phonetic contexts.

### 4.5 Limitations and Future Directions

Several limitations should be considered when interpreting the present findings. Most notably, the number of speakers with abnormal hypernasality was small, particularly at higher severity levels. This imbalance likely reduced statistical power for subgroup analyses and may explain why some correlations were no longer significant when analyses were restricted to speakers with abnormal hypernasality. Future studies should include larger and more balanced samples across the full hypernasality severity spectrum to better characterize severity-dependent effects. Such studies should also examine whether intelligibility-based quality filters, such as WER thresholds, preferentially exclude speakers with more severe speech impairment or hypernasality (Tobin et al., 2024). The MDC estimates were also derived from a relatively small control sample and should be further validated in larger cohorts (de Vet et al., 2006).

A second limitation is that the stop-specific analyses were also based on a limited and uneven set of phonetic contexts. The /p/, /t/, and /k/ tokens differed in number and phonetic context: /p/ occurred twice in different syllabic positions, /t/ occurred twice in syllable-final position and may have been unreleased, and /k/ occurred only once within a consonant cluster. These differences may have contributed to phoneme-specific patterns in NCD and NPP and warrant caution when interpreting differences among stops. Future studies should also examine audible nasal emission during pressure-consonant production and determine how it relates to perceptual hypernasality and consonant-based automated measures such as NPP (Oren et al., 2020).

Another limitation is that hypernasality ratings were based on sentence-level perceptual judgments. Although such ratings reflect current clinical practice, they remain subjective and may be influenced by speech characteristics other than nasal resonance. Future work incorporating complementary instrumental measures of velopharyngeal function may provide additional validation of automated acoustic measures.

Future research should also investigate whether combining NCD and NPP yields improved prediction of hypernasality severity relative to either measure alone. Because the two measures are strongly related but derived from different computational frameworks, their integration may provide complementary information that improves robustness and generalizability across clinical populations.

## 5 Conclusion

The present study demonstrated that both NCD and NPP are sensitive to clinician-rated hypernasality in speakers with MSDs. Consistent with the study hypotheses, increasing hypernasality severity was generally associated with lower NCD values and higher NPP values, although the strength of these relationships varied across stop consonants. NCD and NPP exhibited strong associations with one another and comparable ability to distinguish normal from abnormal hypernasality. Furthermore, hypernasality-related information was observed not only in stop consonants but also across broader classes of speech sounds, with consonant and vowel NPP measures demonstrating similar relationships with severity and equivalent classification performance. Together, these findings support the use of automated acoustic measures as objective markers of hypernasality and suggest that phonological feature-based approaches may provide a flexible framework for assessing nasal resonance across diverse speech contexts.

## Data Availability Statement

The datasets of the current study are available from the corresponding author on reasonable request.

## Acknowledgments

The authors extend gratitude to these patients and their families for their time and dedication to this research program.

## Funding Statement

This work was supported by the grant from the National Institute of Health, National Institute on Aging R01 AG083832 (PI: Botha).

## Conflicts of Interest

The authors have declared that no competing financial or non-financial interests existed at the time of publication.

## Footnotes

1 NCD code: https://github.com/michaelsaxon/ncd

2 Phonet code: https://github.com/jcvasquezc/phonet

## Notes

### Competing Interest Statement

The authors have declared no competing interest.

### Author Declarations

Ethics committee of Mayo Clinic gave ethical approval for this work.

